# Sparse Deep Neural Networks on Imaging Genetics for Schizophrenia Case-Control Classification

**DOI:** 10.1101/2020.06.11.20128975

**Authors:** Jiayu Chen, Xiang Li, Vince D. Calhoun, Jessica A. Turner, Theo G. M. van Erp, Lei Wang, Ole A. Andreassen, Ingrid Agartz, Lars T. Westlye, Erik Jönsson, Judith M. Ford, Daniel H. Mathalon, Fabio Macciardi, Daniel S. O’Leary, Jingyu Liu, Shihao Ji

## Abstract

Machine learning approaches hold potential for deconstructing complex psychiatric traits and yielding biomarkers which have a large potential for clinical application. Particularly, the advancement in deep learning methods has promoted them as highly promising tools for this purpose due to their capability to handle high-dimensional data and automatically extract high-level latent features. However, current proposed approaches for psychiatric classification or prediction using biological data do not allow direct interpretation of original features, which hinders insights into the biological underpinnings and development of biomarkers. In the present study, we introduce a sparse deep neural network (DNN) approach to identify sparse and interpretable features for schizophrenia (SZ) case-control classification. An *L*_0_-norm regularization is implemented on the input layer of the network for sparse feature selection, which can later be interpreted based on importance weights. We applied the proposed approach on a large multi-study cohort (N = 1,684) with brain structural MRI (gray matter volume (GMV)) and genetic (single nucleotide polymorphism (SNP)) data for discrimination of patients with SZ vs. controls. A total of 634 individuals served as training samples, and the resulting classification model was evaluated for generalizability on three independent data sets collected at different sites with different scanning protocols (n = 635, 255 and 160, respectively). We examined the classification power of pure GMV features, as well as combined GMV and SNP features. The performance of the proposed approach was compared with that yielded by an independent component analysis + support vector machine (ICA+SVM) framework. Empirical experiments demonstrated that sparse DNN slightly outperformed ICA+SVM and more effectively fused GMV and SNP features for SZ discrimination. With combined GMV and SNP features, sparse DNN yielded an average classification error rate of 28.98% on external data. The importance weights suggested that the DNN model prioritized to select frontal and superior temporal gyrus for SZ classification when a high sparsity was enforced, and parietal regions were further included with a lower sparsity setting, which strongly echoed previous literature. This is the first attempt to apply an interpretable sparse DNN model to imaging and genetic features for SZ classification with generalizability assessed in a large and multi-study cohort. The results validate the application of the proposed approach to SZ classification, and promise extended utility on other data modalities (e.g. functional and diffusion images) and traits (e.g. continuous scores) which ultimately may result in clinically useful tools.

## Introduction

Schizophrenia (SZ), is a disabling psychiatric disorders with a lifetime prevalence ~0.8%, casts a serious socioeconomic burden worldwide [1]. More than a century after Kraepelin’s dichotomy was formulated, precise treatment is still not available for SZ [2, 3]. Current diagnostic and treatment practice are largely based on descriptive clinical characteristics whose relationships to underlying biological processes await delineation [2, 4]. This gap underlies many issues faced by clinical psychiatry, including vague boundaries between defined clinical entities, and heterogeneity within individual clinical entities. As a result, symptom presentations often do not neatly fit the categorical diagnostic system, and one diagnostic label covers biologically diverse conditions. These issues challenge treatment planning, which turns out to be largely empirical [5, 6]. It has now been widely acknowledged that objective biological markers are needed to quantify abnormalities underlying phenotypic manifestation, which allows characterizing disorders based on a multitude of dimensions and along a spectrum of functioning, so as to improve patient stratification and inform treatment planning [7, 8].

Hopes have been invested in machine learning approaches as a solution to this challenge, given the complexity of SZ. Patients with SZ present widespread structural and functional brain abnormalities, including gray matter loss in the frontal, temporal and parietal cortices and subcortical structures [9–11], reduced fractional anisotropy in most major white matter fasciculi [12], as well as abnormal resting state functional connectivity in default mode, somatomotor, visual, auditory, executive control and attention networks [13–15]. In parallel, genome wide association studies (GWASs) of SZ lend support for a polygenic architecture, where the disease risk is attributable to many genetic variants with modest effect sizes [16]. These findings have boosted the efforts to model SZ in a multivariate framework, which is expected to not only delineate the relationships between individual biomarkers and disease, but also to provide a generalizable mathematical model that can be used to predict risk.

One straightforward approach is to feed voxelwise neurobiological features (e.g. gray matter density) into support vector machine (SVM). With this strategy, Nieuwenhuis et al. obtained a classification accuracy of ~70% which was confirmed in independent data with a sample size of a few hundred [17]. Whether more sophisticated feature selection can be combined with classifiers to yield improved discrimination has also been explored. For instance, resting state connectivity between networks extracted by independent component analysis (ICA), followed by K nearest neighbors, yielded an accuracy of 96% in a data set consisting of 28 controls and 28 patients, which were randomly partitioned to serve as training and testing samples [18]. In addition, fusion of multiple modalities that may carry complementary information of the brain holds promise for further improvement. In a work by Liang et al., combining gray and white matter features resulted in an average classification accuracy of ~76% in 48 controls and 54 patients with first episode SZ, in a 10-fold cross validation set up [19]. In contrast to neurobiological features, genetic variables, such as single nucleotide polymorphisms (SNPs), in general suffer modest effect sizes [16] and could hardly be directly trained for classification. A more commonly used feature for risk discrimination is polygenic risk score (PGRS), which reflects the cumulative risk of multiple variants, and proves to be a generalizable and promising marker for disease discrimination and patient stratification [20, 21], with complementary value for group classification beyond brain MRI and cognitive data [22].

More recently, the advancement of deep learning methods has opened a new perspective on elucidating biological underpinnings of SZ. Deep Neural Networks (DNNs) are known to excel in handling high-dimensional data and automatically identifying high-level latent features, which promotes them as promising tools for better understanding of complex traits such as SZ. In a pioneer study, Plis et al. demonstrated the application of restricted Boltzmann machine-based deep belief network to sMRI data. A classification accuracy of ~90% was obtained with a 10-fold cross validation in 181 controls and 198 patients with SZ [23]. A deep discriminant autoencoder network has been proposed and applied to functional connectivity features, and yielded a leave-site-out classification accuracy of ~81% in 377 controls and 357 patients of SZ [24]. A comparable leave-site-out accuracy of ~80% was observed in 542 controls and 558 patients with SZ, when a multi-scale recurrent neural network was applied to time courses of fMRI data [25]. However, these approaches do not provide importance weights of original biological features indicating their relative contribution to classification, making interpretation less straightforward.

As commonly implemented, DNNs are black-boxes with hundreds of layers of convolution, non-linearities, and gates, optimized solely for competitive performance. While the value of DNN may be backed up with a claimed high accuracy on benchmarks, it would be desired to be able to verify, interpret, and understand the reasoning of the system. This is particularly essential for the psychiatric community, for the purpose of deconstructing complex disorders and facilitating improved treatment. In the current work, we introduce a sparse DNN model which allows identifying sparse and interpretable features for SZ discrimination. The sparsity is achieved with an *L*_0_-norm regularization on the input layer of the network for feature selection. Under the *L*_0_-norm sparsity constraint, the model is trained to select the most important features while retaining the high SZ classification accuracy. We applied the sparse DNN approach on a multi-site gray matter volume (GMV) and SNP data set for SZ discrimination. In brief, a total of 634 individuals (346 controls and 288 patients with SZ) served as the training set, which was internally partitioned for hyperparameter tuning. The resulting classification model was then evaluated for generalizability on three independent data sets (n = 635, 255 and 160, respectively). We examined the classification power of pure GMV features, as well as whether combining GMV with SNP features would benefit classification. The performance of the proposed approach was compared with that yielded by ICA+linear SVM. Empirical experiments demonstrate that the selected voxel regions from sparse DNNs are interpretable and echo many previous neuroscience studies.

## Materials and Methods

### Participants

A total of 1,684 individuals aggregated from multiple studies, including MCIC, COBRE, FBIRN, NU, BSNIP, TOP and HUBIN, were employed for the current study. The institutional review board at each site approved the study and all participants provided written informed consent. Diagnosis of SZ was confirmed using the Structured Clinical Interview for Diagnosis for DSM-IV or DSM-IV-TR. Table 1 provides the primary demographic information of individual study. More details regarding scanning information are listed in Table S1, which also provides a summary of previous publications with description of recruitment. The training sample consisted of 288 cases and 346 controls from MCIC, COBRE, FBIRN and NU. Meanwhile, three independent data sets, BSNIP (n = 635), TOP (n = 255) and HUBIN (n = 160) were used for validation.

**Table 1:** Subject demographic information.

| Cohort | N | Sex (M/F) | Age (mean ± SD) | Age (Min - Max) | Diagnosis (HC/SZ) |
| --- | --- | --- | --- | --- | --- |
| **Training** |  |  |  |  |  |
| MCIC+COBRE+FBIRN+NU | 634 | 459/175 | 35.44 ± 12.12 | 16 - 65 | 346/288 |
| **Validation** |  |  |  |  |  |
| TOP | 255 | 144/111 | 33.75 ± 8.99 | 17 - 62 | 154/101 |
| HUBIN | 160 | 108/52 | 41.69 ± 8.56 | 19 - 56 | 76/84 |
| BSNIP | 635 | 347/287 | 35.95 ± 12.45 | 16 - 64 | 369/266 |

### Structural MRI data

Whole-brain T_1_-weighted images were collected with 1.5T and 3T scanners of various models, as summarized in Table 1 and Table S1. The images of the training set were preprocessed using a standard Statistical Parametric Mapping 12 (SPM12, http://www.fil.ion.ucl.ac.uk/spm) voxel based morphometry pipeline [26–29], a unified model where image registration, bias correction and tissue classification are integrated. The resulting modulated images were resliced to 1.5mm×1.5mm×1.5mm and smoothed by 6mm full width at half-maximum Gaussian kernel. A mask (average GMV > 0.2) was applied to include 429,655 voxels. We further investigated correlations between individual images and the average GMV image across all the subjects. Subjects with correlations < 3SD were considered as outliers and excluded from subsequent analyses [30]. Finally, voxelwise regression was conducted to eliminate the effects from age, sex, and dummy-coded site covariates [28]. While all the scanning parameters (Table S1) would yield 93 dummy variables in the training data, we chose to correct scanning effects by ‘site’ before association analysis to avoid eliminating too much information due to unknown collinearity. The validation images were preprocessed separately, using the same pipeline.

### SNP data

The SNP data were collected and processed as described in our previous work [30]. DNA samples drawn from blood or saliva were genotyped with different platforms (see Table S1). No significant difference was observed in genotyping call rates between blood and saliva samples. A standard pre-imputation quality control (QC) [31] was performed using PLINK [32]. In the imputation, SHAPEIT was used for pre-phasing [33], IMPUTE2 for imputation [34], and the 1000 Genomes data as the reference panel [35]. Only markers with INFO score > 0.3 were retained. Polygenic risk scores (PGRS) for SZ were then computed using PRSice, which was a sum of genetic profiles weighted by the odds ratios reported in the PGC SZ GWAS, reflecting the cumulative risk for SZ of a set of SNPs. Specifically, the genotype data were pruned at r^2^ < 0.1 [30]. Then a full model PGRS was computed on 61,253 SNPs retained after pruning.

### Sparse DNN

Figure 1 shows the overall architecture of our method, which contains three stages. First, the GMV voxels are partitioned into a set of groups (or brain regions) with a pre-defined radius. Then a sparse DNN model is deployed for feature (brain region) selection, followed by augmenting the selected sparse regions of GMV with the SNP data for classifier retraining. In the sequel, we will introduce each of these steps in more details.

**Figure 1:**
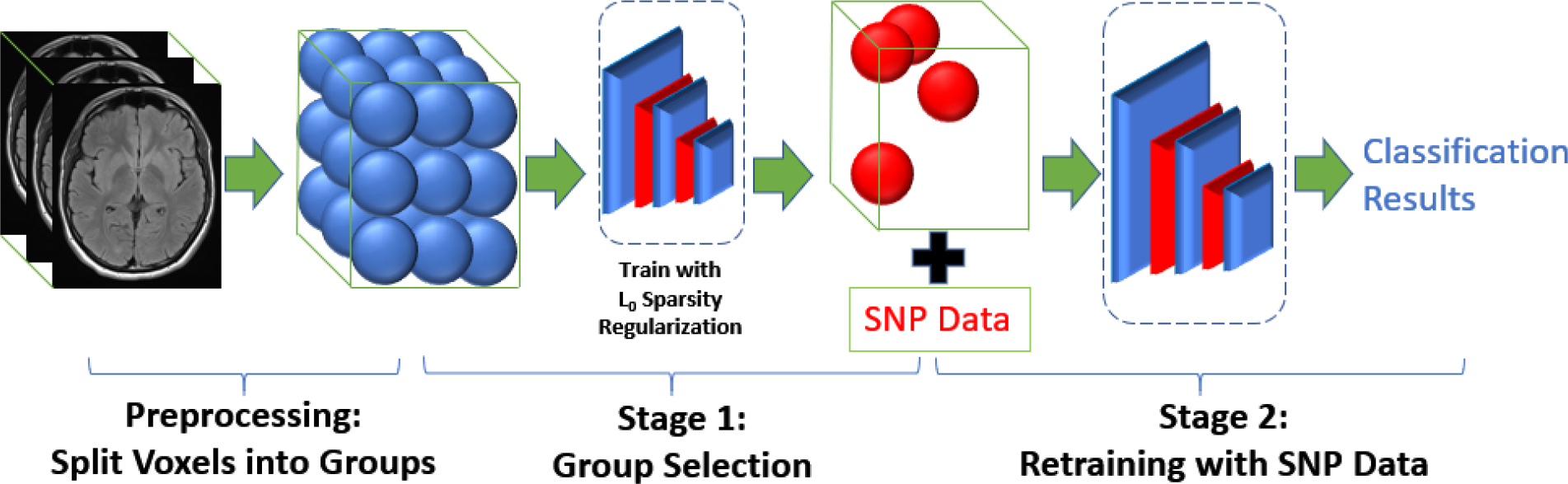
Overall architecture of our method.

Given a GMV dataset *D* = {(***x***_*i*_, *y*_i_), *i* = 1,2, ⋯, *N*}, where ***x***_*i*_ denotes the *i*-th subject’s GMV image and *y*_*i*_ denotes the corresponding label: case or control, we train a neural network *h*(***x***; ***θ***), parameterized by ***θ***, to fit to the dataset *D* with the goal of achieving good generalization to unseen test data. For a GMV image ***x*** ∈ *R*^*M×*1^, we use *x*^*j*^ to represent the *j*-th voxel of image ***x***, where *j* = 1,2, ⋯, *M* and *M* = 429,655 in our study.

As the number of voxels *M* is much larger than the number of functional regions of human brain (e.g., typically around 100 as defined by various brain atlases), we first partition the brain voxels into a set of small regions, each of which is represented by a ball of a pre-defined radius *R*. We enumerate all *M* voxels one by one: if a voxel hasn’t been assigned to any region, we assign that voxel as a root to start a new region. After selecting a root voxel, we compute the Euclidean distance between the root voxel and all the unassigned voxels. All the unassigned voxels with distance smaller than *R* are then assigned into this region. We then iterate this process over the remaining voxels to form next region until all the voxels are assigned to one of the regions. We denote the *k*-th region *G*_*k*_. After this preprocessing step, we identify *K* regions, from which we aim to identify important regions for SZ discrimination.

Stage 1 of our algorithm is to prune insignificant regions from *K* pre-defined regions. We formulate our region selection algorithm by considering a regularized empirical risk minimization procedure with an *L*_0_-norm regularization. Specifically, we attach a binary random variable *z*^*k*^ ∈ {0,1} to all the voxels in region *G*_*k*_:

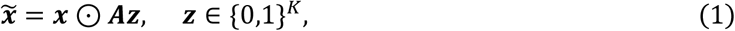

where ***z*** ∈ *R*^*K×*1^ denotes a binary mask for brain image ***x*** ∈ *R*^*M×*1^, ⊙ is an element-wise product, and *A* ∈ *R*^*M×K*^ is an affiliation matrix we construct from the preprocessing step above, with element *A*_*j,k*_ = 1 if voxel *x*^*j*^ is in region *G*_*k*_, and 0 otherwise. For all the voxels in a region *G*_*k*_, they share the same binary mask *z*^*k*^, and *k* ∈ {1,2, ⋯, *K*}. This means if *z*^*k*^ is 0, all the voxels in region *G*_*k*_ will have a value of 0, otherwise the value of *x*^*j*^ is retained. In the sequel, we will discuss our method that can learn ***z*** from training set *D*, and we wish *z*^*k*^ takes value of 1 if *G*_*k*_ is an important region and 0 otherwise. In other words, **z** is a measure of feature (region) importance that we wish to learn from data.

We regard **z** as the feature importance weight for the prediction of DNN model *h*(*x*^*i*^; ***θ***) and learn **z** by minimizing the following *L*_0_-norm regularized loss function:

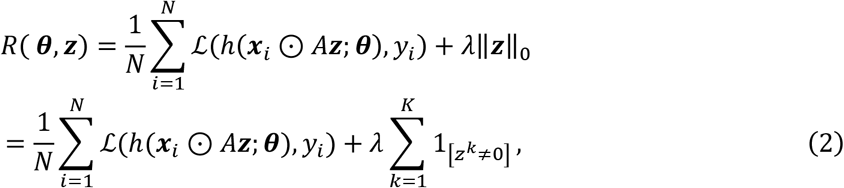

where ℒ(⋅) denotes the data loss over training data *D*, such as the cross-entropy loss for classification, ‖***z***‖_0_ is the *L*_0_-norm that measures number of nonzero elements in ***z***, *λ* is a regularization hyperparameter that balances between data loss and feature sparsity, and 1_[*c*_]is an indicator function that is 1 if the condition c is satisfied, and 0 otherwise. To optimize Eq. 2, however, we note that both the first term and the second term of Eq. 2 are not differentiable w.r.t. **z**. Therefore, further approximations need to be considered.

We can approximate this optimization problem via an inequality from stochastic variational optimization [36]. Specifically, given any function *ℱ*(***z***) and any distribution *q*(***z***), the following inequality holds

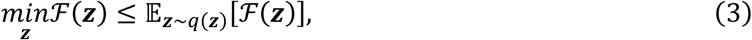

i.e., the minimum of a function is upper bounded by the expectation of the function. With this result, we can derive an upper bound of Eq. 2 as follows.

Since *z*^*k*^, ∀*k* ∈ {1, ⋯, *K*} is a binary random variable, we assume *z*^*k*^ is subject to a Bernoulli distribution with parameter *π*^*k*^ ∈ [0,1], i.e. *z*^*k*^ *~* Ber(*z*; *π*^*k*^). Thus, we can upper bound min *R*(***θ, z***) by the expectation

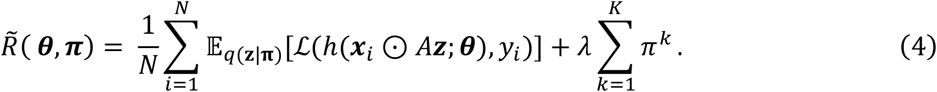

Now the second term of the Eq. 4 is differentiable w.r.t. the new model parameters ***π***. However, the first term is still problematic since the expectation over a large number of binary random variables **z** ∈ {0,1}^*K*^ is intractable, so is its gradient. To solve this problem, we adopt the hard-concrete estimator [37]. Specifically, the hard-concrete gradient estimator employs a reparameterization trick to approximate the original optimization problem of Eq. 4 by a close surrogate loss function

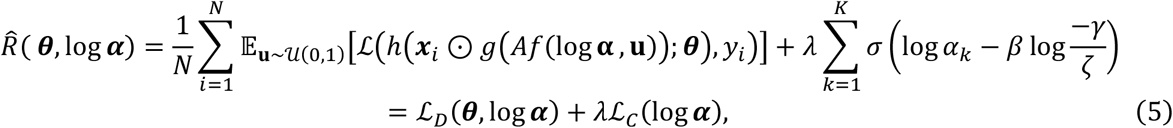

with

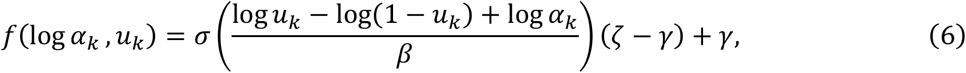

and

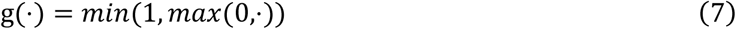

where *σ*(*t*) = 1/(1 + exp (−*t*)) is the sigmoid function, ℒ_*D*_ measures how well the classifier fits to training data *D*, ℒ_*C*_ measures the expected number of non-zeros in ***z***, and 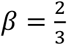, *γ* = −0.1 and *ζ* = 1.1 are the typical parameters of the hard-concrete distribution. Function *g*(*⋅*) is a hard-sigmoid function that bounds the stretched concrete distribution between 0 and 1. With this reparameterization, the surrogate loss function Eq. 5 is differentiable w.r.t. its parameters.

After training, we learn log***α*** from the dataset *D*. At test time, we employ the following estimator to generate a sparse mask or feature importance weight:

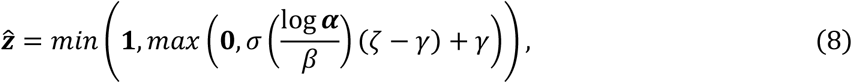

which is the sample mean of ***z*** under the hard-concrete distribution *q*(***z***|log***α***).

After we train the sparse DNN with the *L*_0_-norm regularization, we get the trained neural network parameters ***θ*** and sparse mask 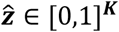 over all *K* regions, with element 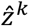 a continuous variable that represents the importance of region *G*_*k*_. Because of the sparsity inducing property of the *L*_0_-norm, many elements of learned 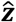 are pushed to zero, which are considered as unimportant regions and thus pruned from the model. The level of sparsity can be modulated by hyperparameter *λ*: the larger *λ* is, the sparser regions is identified, and *vis-a-versa*.

In Stage 2 of our algorithm, we can further improve the accuracy of the classifier by finetuning the DNN with the selected *L* regions from Stage 1 but without the *L*_0_-norm regularization. To examine whether incorporating genetic features can improve the classification accuracy, we also concatenate the PGRS feature to the selected voxels as the input of the DNN classifier to finetune the classifier.

In our study, the training data consists of 634 individuals (346 controls and 288 cases), which were equally partitioned into three subsets (each containing 33% of the samples). A nested 3-fold cross validation was then implemented to identify the discriminating genetic and brain MRI features and construct a classification model for SZ. The region radius *R* we used was 12mm and each brain image was partitioned into 1111 regions as we described above. In Stage 1 group selection and Stage 2 retraining, we used a DNN classifier with 2 fully connected layers of 200 and 16 neurons, respectively, and the rectified linear unit (ReLU) activation function. We performed grid search to find the best hyperparameters for our sparse DNN model. In Stage 1 group selection, we used the SGD optimizer with learning rates of 0.005 and 1 for model parameter ***θ*** and log***α***, respectively. In Stage 2 retraining classifier, we used the Adam optimizer with learning rate of 0.005 for ***θ*** and a weight decay of 1e-5. After the sparse DNN was trained on the GMV features, the regions with nonzero 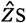s were considered as important regions for the SZ classification. The selected regions across 3-fold cross validation were highlighted for model interpretation. In particular, we tuned hyperparameter *λ* to compare the classification performances with different levels of sparsity, i.e. with 5 or 20 regions as predictors. In Stage 2 retraining, the selected voxel regions were fed into the classifier and may concatenate the PGRS feature to improve the classification accuracy. The model established in the training data was further evaluated on three external data sets: BSNIP, TOP and HUBIN.

### ICA+linear SVM

To compare with sparse DNN, we also conducted classification using linear SVM with components extracted by ICA as input. ICA decomposes data into a linear combination of underlying components among which independence is maximized [38, 39]. When applied to sMRI data, ICA essentially identifies maximally independent components, each including a weighted pattern of voxels with covarying gray matter patterns across samples [40]. ICA has been widely used in the neuroimaging field, yielding meaningful and generalizable brain networks which are not well captured by anatomical atlas [41, 42, 28]. In the current work, following the training and testing of the sparse DNN, we applied ICA on the GMV data for 67% of the training samples. The resulting components were then fed into linear SVM to obtain a classification model. This model was then assessed on the remaining 33% of the training samples for accuracy. Since the number of ICA components was a hyperparameter to be tuned, we repeated the above process with different component numbers. The optimal model was then determined to be the one yielding the highest accuracy, and this was then validated in the three independent data sets. Echoing the sparse DNN experiments, we also investigated whether having more GMV components as predictors would affect the performance of classification. When genetic feature was further incorporated, PGRS was treated as an additional predictor, which was sent into linear SVM along with the GMV components. Note that genetic data were available only for TOP and HUBIN, such that only these two data sets were examined for imaging genetic based classification.

## Results

The performance was summarized in Table 2. When only GMV features were used for classification, the ICA+SVM approach achieved the highest accuracy with 20 components in the training samples. In parallel, the performance of DNN also started to saturate around a sparsity level of 20 regions. It can be seen that for both ICA and DNN approaches, lower error rates were achieved when 20 rather than only 5 brain regions/components served as predictors. When fewer brain regions were used to train the model, the mean error rate across three independent data sets was ~35% for both ICA and DNN, though in specific data sets discrepancies could be noted. When the classification model was allowed to incorporate more brain regions/components, the mean error rate across three data sets decreased to 31.03% for DNN models and 31.86% for ICA models. Specifically, the error rates were comparable between ICA and DNN in HUBIN and BSNIP, while the error rate improved by 3.66% in TOP when DNN was used.

**Table 2:** Summary of classification error rates.

|  | sMRI |  |  | sMRI + SNP |  |
| --- | --- | --- | --- | --- | --- |
|  | TOP (255) | HUBIN (160) | BSNIP (635) | TOP (255) | HUBIN (160) |
| DNN (5 regions) |  |  |  |  |  |
| EER1 | 35.69 | 33.08 | 34.49 | 32.94 | 28.13 |
| EER2 | 34.90 | 36.25 | 33.60 | 33.33 | 33.13 |
| EER3 | 34.90 | 36.25 | 36.80 | 32.55 | 32.5 |
| EER mean | 35.16 | 35.19 | 34.96 | 32.94 | **31.25** |
| ICA+SVM (5 ICs) |  |  |  |  |  |
| EER1 | 36.86 | 31.88 | 37.17 | 30.20 | 35.00 |
| EER2 | 37.25 | 34.38 | 37.32 | 30.59 | 35.63 |
| EER3 | 34.90 | 32.50 | 36.85 | 29.80 | 35.63 |
| EER mean | 36.34 | 32.92 | 37.11 | **30.20** | 35.42 |
| DNN (20 regions) |  |  |  |  |  |
| EER1 | 30.59 | 28.13 | 31.16 | 30.65 | 26.27 |
| EER2 | 30.98 | 32.50 | 32.91 | 27.75 | 27.25 |
| EER3 | 33.33 | 28.75 | 31.02 | 32.26 | 28.24 |
| EER mean | 31.63 | 29.79 | 31.69 | **30.22** | **27.75** |
| ICA+SVM (20 ICs) |  |  |  |  |  |
| EER1 | 33.33 | 27.50 | 30.87 | 32.94 | 29.38 |
| EER2 | 39.22 | 31.25 | 31.02 | 35.29 | 33.75 |
| EER3 | 33.33 | 28.75 | 31.50 | 30.98 | 30.00 |
| EER mean | 35.29 | 29.17 | 31.13 | 33.07 | 31.04 |

When PGRS was further incorporated for classification, the DNN approach yielded consistent improvement in accuracy across all the data sets, either with 5 or 20 regions as predictors, where the decrease in error rate ranged from 1.41% to 3.94%. In contrast, with ICA components were combined with PGRS for classification, the error rate did not always decrease. The lowest error rate (27.75%) was observed in HUBIN, when the DNN classification model used 20 brain regions plus the PGRS.

The brain regions identified by DNN are summarized in Tables 3 (5 regions) and 4 (20 regions), and Figures 2 and 3 show the spatial maps of individual regions. Note that only the regions identified in all three folds are listed. When 5 regions were to be selected as predictors, the three folds consistently identified the same 5 regions, spanning inferior, middle and superior frontal gyrus, superior temporal gyrus, as well as cerebellum. When 20 regions were to be selected, variations were noted across folds, such that 13 brain regions were consistently identified. Compared to those covered by 5 regions, cuneus, precuneus, medial frontal gyrus, and paracentral lobule were also determined to be informative and included for classification. The importance weights yielded by the interpretable DNN model were overall highly consistent with those inferred from the original features, such that a positive/negative DNN weight indicated that the region showed higher/lower values in controls compared to patients with SZ. The only exception was region 27 which was identified in the 20-region model.

**Table 3:**
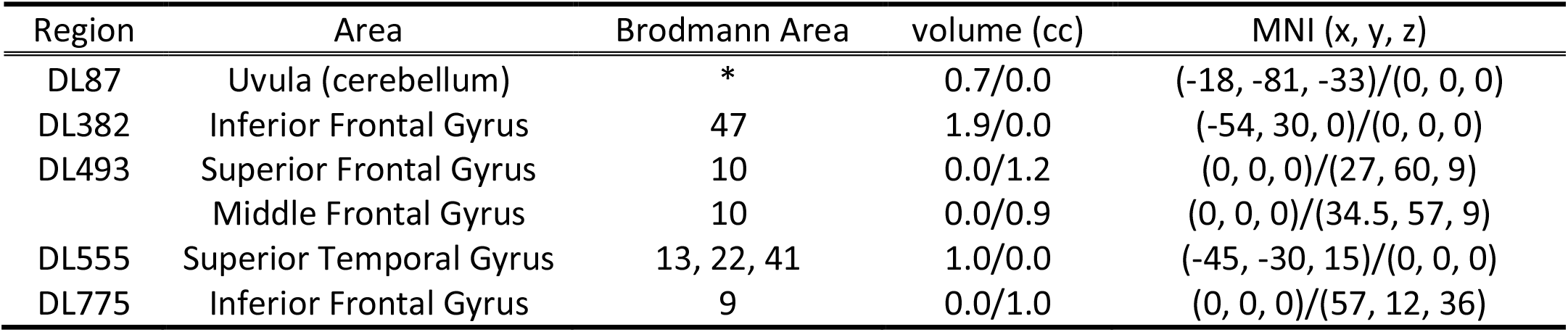
Summary of the 5 important brain regions identified by DNN.

| Region | Area | Brodmann Area | volume (cc) | MNI (x, y, z) |
| --- | --- | --- | --- | --- |
| DL87 | Uvula (cerebellum) | * | 0.7/0.0 | (-18, -81, -33)/(0, 0, 0) |
| DL382 | Inferior Frontal Gyrus | 47 | 1.9/0.0 | (-54, 30, 0)/(0, 0, 0) |
| DL493 | Superior Frontal Gyrus | 10 | 0.0/1.2 | (0, 0, 0)/(27, 60, 9) |
|  | Middle Frontal Gyrus | 10 | 0.0/0.9 | (0, 0, 0)/(34.5, 57, 9) |
| DL555 | Superior Temporal Gyrus | 13, 22, 41 | 1.0/0.0 | (-45, -30, 15)/(0, 0, 0) |
| DL775 | Inferior Frontal Gyrus | 9 | 0.0/1.0 | (0, 0, 0)/(57, 12, 36) |

**Figure 2:**
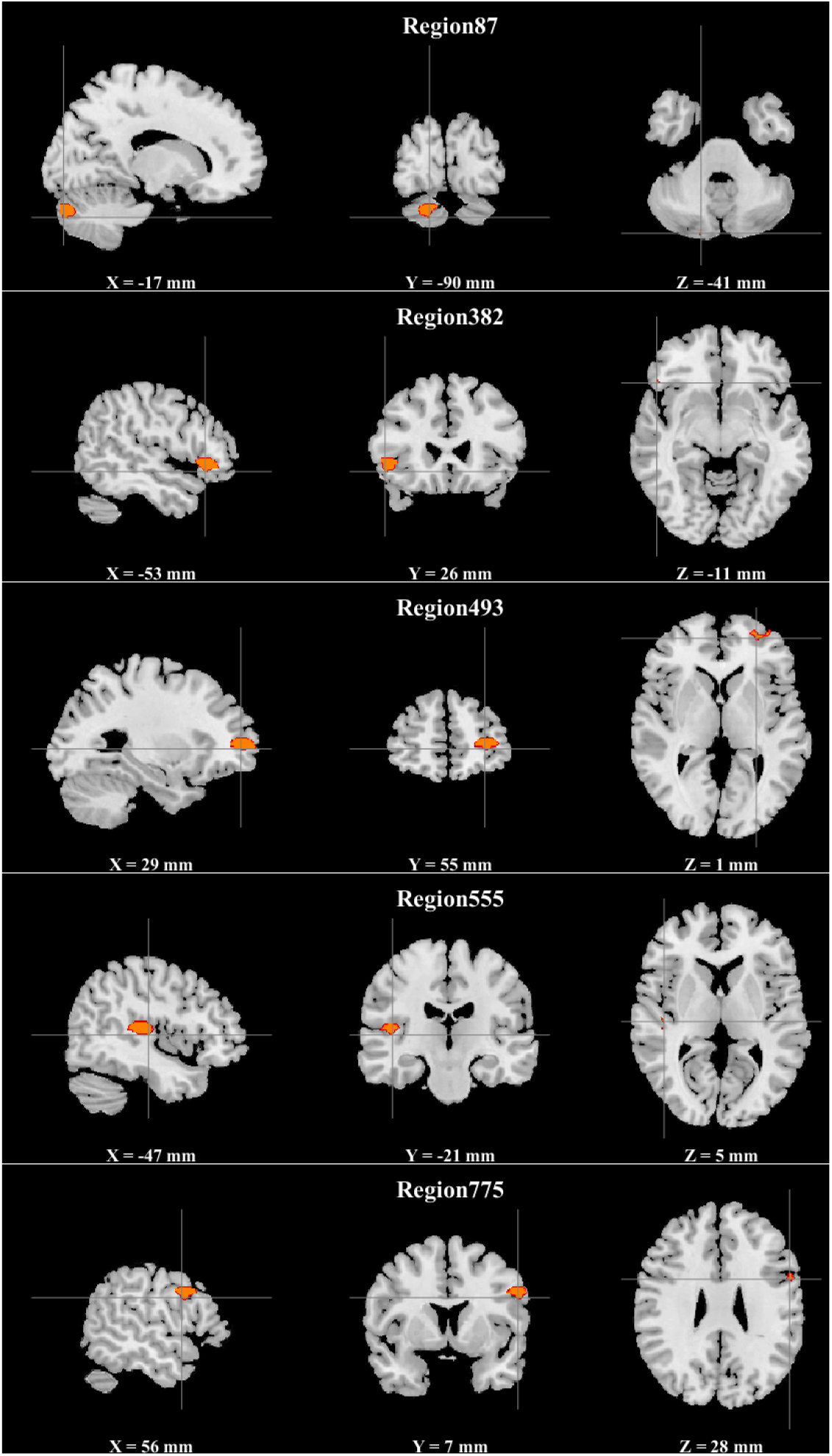
Spatial maps of the five schizophrenia-discriminating regions identified by sparse DNN.

**Figure 3:**
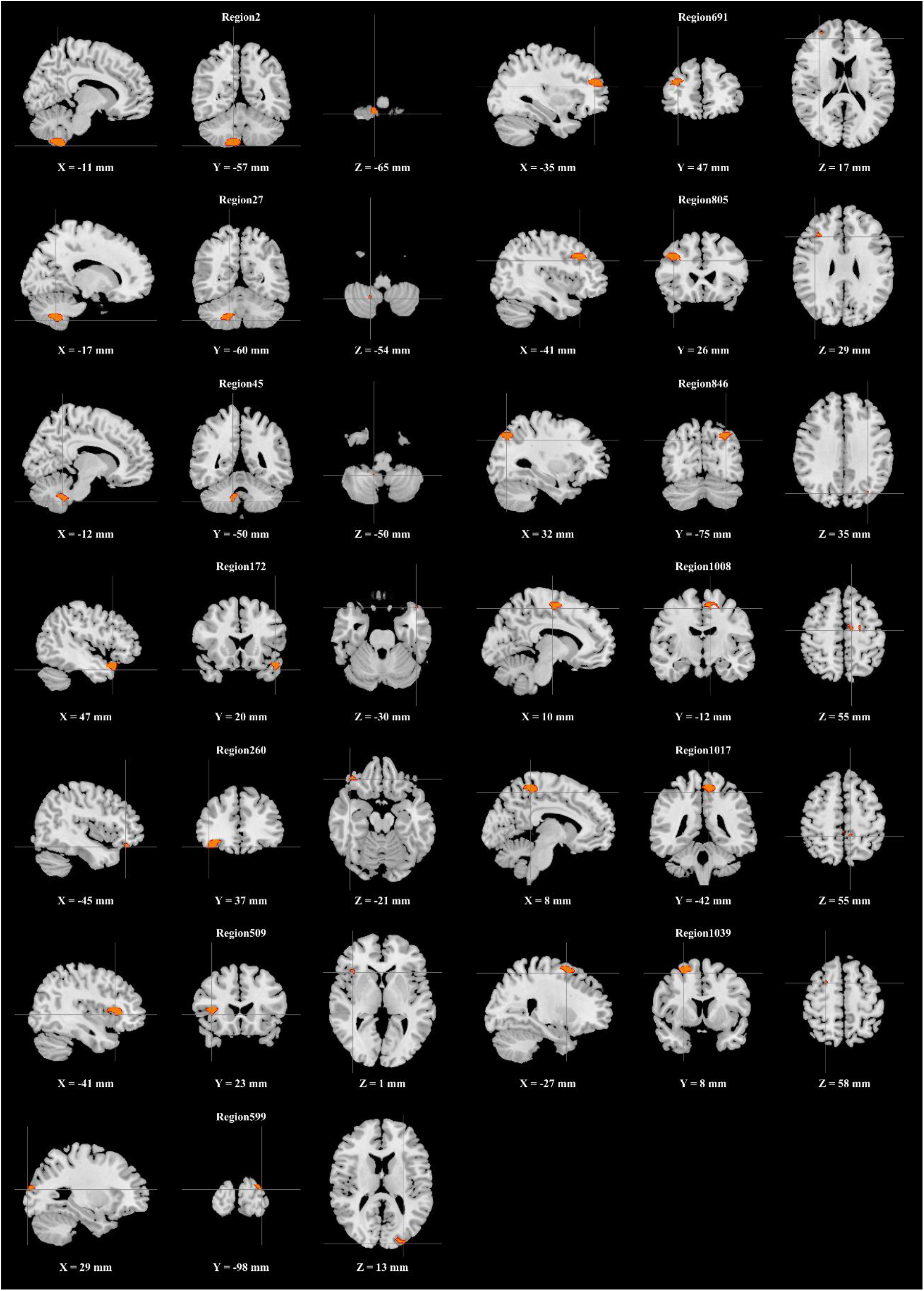
Spatial maps of the 13 schizophrenia-discriminating regions identified by sparse DNN.

## Discussion

An interpretable sparse DNN approach was proposed for application to medical data analysis and its capability was examined on a large and heterogeneous SZ data set. The results confirmed that the proposed approach yielded reasonable classification accuracies, could identify meaningful brain regions, and the interpretation of these brain regions was consistent with that directly inferred from original features. Particularly, the proposed model appeared to more effectively fuse imaging and genetic features for classification compared to ICA+SVM, holding potential for data fusion.

The DNN models reliably generalized to data collected at different sites, with reasonable classification accuracies compared to ICA+SVM. The generalizability indicates that the classification models are not vulnerable to scanning protocol, recruiting criteria, ethnicity influence, medication history, etc. Regarding performance, both DNN and ICA+SVM approaches presented higher accuracies when more brain regions/components served as predictors, with error rates being 31.03% and 31.86%, respectively. The ICA+SVM performance was comparable to those reported by Cai et al., where the authors conducted a comprehensive study on generalizability of machine learning for SZ classification using ICA-extracted resting-state fMRI features, and achieved an external accuracy of 70% with transfer learning procedures [43]. Notably, Cai et al. emphasized the importance of assessing models across sites and studies, while results based on a single study need to be interpreted cautiously. This might explain why our classification accuracy based on a large and multi-study cohort is lower than some previous studies with smaller sample sizes or single-study cohort [44], indicating complex heterogeneity of patients with SZ. Increasing sample size of the training data and incorporating other data modalities promise further improvement.

The proposed approach highlights a sparsity constraint, which allows trade-off between explained variance and interpretability of identified features. In general, a low level of sparsity allows more features to be admitted into the classification model, which however results in more variance across samples. As shown in the current work, when a higher level of sparsity was enforced, the same 5 regions were identified across 3 folds. In contrast, with a lower sparsity, 13 out of 20 regions were consistently identified, although the latter explained more variance and yielded higher classification accuracies. It should be pointed out that, increasing the predictors from 5 to 20 regions resulted in a decrease of ~4% in error rate, which was indeed not profound. In other words, although GMV abnormalities are widely distributed across the brain in SZ, the majority of the variance can be captured by the identified five distinct regions. The samples missed in the classification, or missing variance, likely call for a larger training data set to allow better capturing heterogeneity, as well as for information from other data modalities, rather than simply adding more features from the sMRI modality.

SZ is a complex disorder, where genetic and environmental factors interact with each other to affect brain structure and function which ultimately manifest into clinical symptoms. With so many factors involved in the pathology of SZ, it is expected that multiple data modalities need to be integrated to fully characterize the disorder. This also applies to classification, which should capitalize on data fusion to extract complementary information from different modalities. The proposed model holds promise for this purpose. In all the tested scenarios, the DNN approach effectively fused GMV and PGRS features to yield improved classification accuracies, indicating that the model reliably extracted SZ-related variance in PGRS that was not captured by GMV. In contrast, no consistent improvement was noted for ICA+SVM, where PGRS and brain components were directly fed into linear SVM for classification training. The results appeared to lend support that nonlinear models excel in delineating the relationships across different modalities in hidden layers and robustly capturing complementary variance that is related to the trait of interest.

The brain regions identified by DNN are overall well documented in SZ studies. With high sparsity, 5 brain regions were consistently identified across 3 folds, as listed in Table 3, highlighting frontal gyrus, superior temporal gyrus, and cerebellum. All the five regions presented positive weights, indicating higher GMV in controls compared to patients, which was consistent with the results of two-sample t-tests on original GMV features. SZ-related gray matter reduction has been widely observed in temporal and frontal regions. A longitudinal study by Thompson et al. revealed accelerated gray matter loss in early-onset SZ, with earliest deficits found in parietal regions and progressing anteriorly into temporal and prefrontal regions over 5 years [45]. The identified frontal and temporal brain regions have also been identified for SZ-related reduction in a comprehensive study on gray matter volume in psychosis using the BSNIP cohort [9]. The role of cerebellum in SZ has been revised in recent years, where accumulating evidence suggests that cerebellum is also involved in cognitive functions and cerebellar abnormalities are noted in SZ [46, 47]. Gray matter loss around the identified cerebellar region has also been reported previously [48].

With low sparsity, 13 brain regions were consistently identified by DNN across 3 folds, as listed in Table 4. In addition to frontal, temporal and cerebellar regions discussed above, parietal regions including cuneus, precuneus and paracentral lobule were highlighted. As implicated in Thompson et al, while temporal and prefrontal gray matter loss were characteristic of adult SZ, parietal regions were noted for earliest gray matter loss which was faster in younger patients with SZ [45]. The identified parietal regions also echoed the BSNIP findings to show higher GMV in controls compared to patients [9]. Overall, it is reasonable that DNN prioritized to select temporal and frontal regions for classification when high sparsity was enforced, which aligns with the notion that gray matter loss in these regions characterizes adult SZ. In the meantime, when a lower sparsity was enforced, parietal abnormalities were the first priority to be added as additional predictors which offered complementary variance. Among the 13 regions, region 27 (cerebellar tonsil) was the only feature whose DNN weights did not coincide with the inference drawn from original GMV features. It was noted that the voxels in region 27 showed modest case-control differences compared to voxels in other identified brain regions. We suspect the selection of region 27 by DNN might be driven by some hidden properties rather than group differences, which explains the inconsistency in interpretation between DNN and two-sample t-tests.

**Table 4:** Summary of the 13 important brain regions identified by sparse DNN.

| Region | Area | Brodmann Area | volume (cc) | MNI (x, y, z) |
| --- | --- | --- | --- | --- |
| DL2 | Inferior Semi-Lunar Lobule | * | 0.1/0.0 | (-7.5, -60, -54)/(0, 0, 0) |
| DL27 | Cerebellar Tonsil | * | 1.4/0.0 | (-15, -55.5, -43.5)/(0, 0, 0) |
| DL45 | Cerebellar Tonsil | * | 0.7/0.0 | (-12, -55.5, -40.5)/(0, 0, 0) |
| DL172 | Superior Temporal Gyrus | 38 | 0.0/1.0 | (0, 0, 0)/(48, 22.5, -19.5) |
| DL260 | Middle Frontal Gyrus | 11 | 0.9/0.0 | (-37.5, 40.5, -10.5)/(0, 0, 0) |
| DL509 | Inferior Frontal Gyrus | 13, 47 | 1.3/0.0 | (-42, 25.5, 10.5)/(0, 0, 0) |
| DL599 | Cuneus | 18, 19 | 0.0/1.0 | (0, 0, 0)/(18, -88.5, 19.5) |
| DL691 | Middle Frontal Gyrus | 10, 46 | 1.2/0.0 | (-34.5, 46.5, 27)/(0, 0, 0) |
| DL805 | Middle Frontal Gyrus | 9 | 2.0/0.0 | (-45, 28.5, 39)/(0, 0, 0) |
| DL846 | Precuneus | 7, 19 | 0.0/1.0 | (0, 0, 0)/(30, -66, 42) |
| DL1008 | Medial Frontal Gyrus | 6 | 0.0/1.3 | (0, 0, 0)/(7.5, -4.5, 63) |
| DL1017 | Paracentral Lobule | 4, 5, 6 | 0.2/1.7 | (-1.5, -40.5, 61.5)/(4.5, -37.5, 64.5) |
| DL1039 | Middle Frontal Gyrus | 6 | 1.3/0.0 | (-21, 9, 67.5)/(0, 0, 0) |

One limitation of our algorithm is that we assume the brain regions to be spherical, which we obtained by measuring the Euclidean distance. This may not align with the optimal partition. And we did not extensively investigate how the radius of brain regions would affect the performance. In the future, we plan to test whether defining regions based on a brain atlas (such as Yeo atlas [49]) would benefit the model training. Besides, likely due to the limited sample size, the DNN performance saturated at 2 hidden layers. It remains a question how the performance would scale with increasing sample size. This awaits investigation when more data become available. Furthermore, while the DNN approach holds promise for data fusion, its capability of integrating multiple high-dimensional imaging modalities was not examined in the current work, given that incorporating another modality would further reduce the sample size. This will also be part of our future work.

In summary, to the best of our knowledge, this is the first study of DNN application to sMRI and genetic features for SZ case-control classification with generalizability assessed in a large and multi-study cohort. An interpretable sparse DNN approach was first proposed to allow identifying, refining and interpreting features used in classification. The results indicate that the new approach yielded reasonable classification performances, highly interpretable classification features, as well as potential for data fusion. Collectively, the current work validates the application of the proposed approach to SZ classification, and promises extended utility on other data modalities (e.g. functional and diffusion images) and traits (e.g. continuous scores).

## Data Availability

The MCIC and COBRE data are available through COINS (https://coins.mrn.org). The NU imaging data can be accessed through SchizConnect (http://schizconnect.org/) and the BSNIP imaging data through NIMH Data Archive (https://nda.nih.gov/). Request of access to other data should be addressed to the individual principal investigator.

## Acknowledgements

This project was funded by the National Institutes of Health (P20GM103472, P30GM122734, R01EB005846, 1R01EB006841, R01MH106655, 5R01MH094524, U24 RR021992, U24 RR025736-01, U01 MH097435, R01 MH084803, R01 EB020062), National Science Foundation (1539067, 1636893, 1734853), Research Council of Norway (RCN#223273), K. G. Jebsen Stiftelsen and South-East Norway Health Authority.

